# Covid-19 lockdown: Ethnic differences in children’s self-reported physical activity and the importance of leaving the home environment. A longitudinal and cross-sectional study from the Born in Bradford birth cohort study

**DOI:** 10.1101/2021.02.26.21252543

**Authors:** Daniel D Bingham, Andy Daly-Smith, Jennifer Hall, Amanda Seims, Sufyan A. Dogra, Stuart J Fairclough, Mildred Ajebon, Brian Kelly, Bo Hou, Katy A Shire, Kirsty L Crossley, Mark Mon-Williams, John Wright, Kate Pickett, Rosemary McEachan, Josie Dickerson, Sally E Barber, on behalf of the Bradford Institute for Health Research COVID-19 Scientific Advisory Group

**Affiliations:** Bradford Institute for Health Research, Bradford Teaching Hospitals NHS Foundation Trust, Bradford Royal Infirmary, Duckworth Lane, Bradford, BD9 6RJ, UK; Faculties of Life Sciences and Health Studies, University of Bradford, Richmond Road, Bradford, BD7 1DP, UK; Centre for Applied Education Research, Wolfson Centre for Applied Health Research, Bradford Royal Infirmary, West Yorkshire, UK; Health Research Institute and Movement Behaviours, Health, and Wellbeing Research Group, Dept. Sport and Physical Activity, Edge Hill University, Ormskirk, L39 4QP, UK; Department of Health Sciences, University of York, Seebohm Rowntree Building, University of York, Heslington, York, YO10 5DD, UK; School of Psychology, University of Leeds, Leeds LS2 9JT

**Keywords:** COVID-19, lockdown, physical activity, children, ethnicity, moderate-to-vigorous, self-report, correlates, environment

## Abstract

**Background:** In England, the onset of COVID-19 and a rapidly increasing infection rate resulted in a lockdown (March-June 2020) which placed strict restrictions on movement of the public, including children. Using data collected from children living in a multi-ethnic city with high levels of deprivation, this study aimed to: (1) report childrens self-reported physical activity (PA) during the first COVID-19 UK lockdown and identify associated factors; (2) examine changes of childrens self-reported PA prior to and during the first UK lockdown.

**Methods:** This study is part of the Born in Bradford (BiB) COVID-19 Research Study. PA (amended Youth Activity Profile), sleep, sedentary behaviours, daily frequency/time/destination/activity when leaving the home, were self-reported by 949 children (9-13 years). A sub-sample (n=634) also self-reported PA (Physical Activity Questionnaire for Children) pre-pandemic (2017-February 2020). Univariate analysis assessed differences in PA between sex and ethnicity groups; multivariable logistic regression identified factors associated with children’s PA. Differences in children’s levels of being sufficiently active were examined using the McNemar test examined change in PA prior to and during the lockdown, and multivariable logistic regression to identify factors explaining change.

**Results:** During the pandemic, White British (WB) children were more sufficiently active (34.1%) compared to Pakistani Heritage children (PH) (22.8%) or ‘Other’ ethnicity children (O) (22.8%). WB children reported leaving the home more frequently and for longer periods than PH and O children. Modifiable variables related to being sufficiently active were frequency, duration, type of activity, and destination away from the home environment. There was a large reduction in children being sufficiently active during the first COVID-19 lockdown (28.9%) compared to pre-pandemic (69.4%).

**Conclusions:** Promoting safe extended periods of PA everyday outdoors is important for all children, in particular for children from ethnic minority groups. Children’s PA during the first COVID-19 UK lockdown has drastically reduced from before. Policy and decision makers, and practitioners should consider the findings in order to begin to understand the impact and consequences that COVID-19 has had upon children’s PA which is a key and vital behaviour for health and development.

## Background

In England, the immediate response to the first wave of Severe Acute Respiratory Syndrome Coronavirus 2 (SARS-CoV-2) - COVID19 - pandemic was a stringent lockdown implemented on 23^rd^ March 2020.^1^ The government placed extreme restrictions on movement of the public stating that *“during the emergency period, no person may leave the place where they are living without reasonable excuse”,* which included shopping for food and medical supplies.^1^ Furthermore, guidance stipulated that members of the public could also leave the home for a short bout (60 mins) of local daily exercise. All playgrounds and indoor and outdoor play facilities (e.g. skate parks, soft play centres) were closed, in addition to leisure facilities and gyms. Schools were closed for most children with the exception of vulnerable children and children of key workers (those working across health, social and public sectors). The lockdown measures were eased in England on 4^th^ July 2020.^2^ However, at the time of writing, two further national lockdowns have occurred in England, in November-2020^3^ and January 2021 (currently ongoing).^3, 4^

Before the COVID-19 pandemic, national and international epidemiological data (whether device or self-reported measured) report that up to 80% of children and young people in high-income countries are not sufficiently physically active for health and well-being (e.g. achieve 60 minutes of MVPA per day).^5, 6^ Within England, recent survey data suggests that the 53.2% of children, aged 5-16 years, were not achieving physical activity (PA) guidelines.^7^ Of specific concern, levels of inactivity were higher in children from ethnic minority groups, especially those with South Asian heritage.^8, 9^ Such low levels of PA place children at risk of poor physical and mental wellbeing in addition to having a negative impact on school performance.^10–14^Within South Asian communities, such risks are high as children present with increased rates of Obesity and type-II diabetes.^15^ Further, early evidence suggest such populations are more likely suffer the most during and after the pandemic.^4, 16–20^ Research conducted during COVID-19 has already reported low levels and significant reductions of children’s PA.^21, 22^ It is essential to understand the impact of the pandemic on PA levels and behaviours for different ethnic groups for two reasons, first to prevent inactive behaviours becoming entrenched and second, to tailor support for different populations by addressing the root causes of PA inequality within different populations.^23^

The Born in Bradford (BiB) research programme^24^ provides a premium opportunity to study the impact of COVID-19 lockdown on school-aged children living in a deprived and ethnically diverse city. To date, BiB has tracked/monitored the health, wellbeing, and determinants of health of over 30,000 Bradford residents (parents and children) since 2007.^24^ The latest round of data collection occurred pre COVID (2017-March 2020, n=7500, aged between 6-11 years)^25^, establishing a pre-COVID baseline; providing a unique opportunity to understand the impact of the COVID lockdown on physical activity behaviour in an ethnically diverse sample of school-aged children and young people. Further, the BiB cohort study will follow participants throughout the duration of the pandemic and in the following years, to understand the impact of the crisis on health and wellbeing trajectories.^26^

The current study is part of the wider Born in Bradford COVID-19 Research Study^26^ and aims to: 1) report childrens self-reported physical activity (PA) during the first COVID-19 UK lockdown and identify associated factors; 2) examine changes of childrens self-reported PA prior to and during the first UK lockdown.

## Methods

### Setting

Bradford is the fifth largest metropolitan district in England with a population of 530,000.^24^ It is a ethnically diverse city situated in the North of England, with almost half of the births in the city are to women of South Asian (mostly Pakistani) heritage.^27–29^ Levels of poverty and ill health (including cardiovascular disease and diabetes) in Bradford are some of the highest in England, and a large proportion of households are classed as overcrowded.^28^ Almost a quarter of Bradford children live in poverty and 24% are living with obesity at age 11 while the rates of childhood obesity are 10% higher among same age group of South Asian children.^28^ Such socio-economic and structural characteristics of Bradford make the community particularly vulnerable to COVID-19.

### Participants and procedure

Participants were children aged 9-13 who were invited to take part in the BiB COVID-19 research study^26^ following a protocol approved by the Health Research Authority and Bradford/Leeds research ethics committee (reference: 16/YH/0320).

The parents/carers of 5,298 children aged 9-13 years who are participants in the existing BiB birth cohort study and who had engaged in a recent follow-up data collection wave pre-COVID-19 (2017-early March 2020)^25^, were contacted by trained researchers via telephone to invite their child to take part in a survey. Following verbal consent from parents/carers, children received a survey via post to be completed and returned to the research team using pre-paid envelopes.^26^ Completion of the survey was deemed as participation assent from the child. Overall, 970 children returned surveys during the period of May 21st to July 31st 2020 and 949 children (97.8%) provided enough data to be included for the analysis of the first two aims of this research. The most recent BiB follow-up^25^ included PA survey data for 634 (23.6%) of these 949 children and was used for the analysis of the second aim.

### Measures

#### Demographic measures

Children’s age, sex, ethnicity, and home postcode-derived Index of Multiple Deprivation (IMD)^29^ were extracted from the BiB cohort dataset. Three categories of ethnicity were used for the analysis, White British (WB), Pakistani Heritage (PH) (the two largest groups in the sample) and ‘Other’ (O) (any other ethnic group). Index of Multiple Deprivation (IMD) deciles^30^ were categorised into either the ‘most deprived nationally’ (most deprived 10% areas in England), ‘2^nd^ most deprived nationally’ (10-20% most deprived areas), ‘3^rd^ most deprived nationally’ (30-40% most deprived areas), and ‘4^th^ or more most deprived nationally’ (40%-100% most deprived areas). Child’s school attendance during the April-June 2020 lockdown period was included in the survey.

#### During COVID-19 Lockdown

During the first COVID-19 PA, sedentary behaviours, screen-time, sleep, activity (frequency, duration, type, and place) away from home environment were all measured by child self-report (Table 1). Self-reported PA was measured using a modified version of the validated seven day recall questionnaire, the Youth Activity Profile-English Youth Version (YAP).^31, 32^ The YAP requires children to report the frequency and/or duration of physical activities through different segments of a usual day (i.e. before school, break time at school, lunch at school, after school). During the first lockdown most children were not attending school, so this format was not appropriate, and for the same reason, neither was the PA questionnaire for children (PAQ-C)^33^ which was the questionnaire completed by children in data collection pre-COVID.^25^ The choice to amend the YAP, which was originally based upon the PAQ-C, and to not use the PAQ-C was due to the YAP specifically including an item asking directly for an estimation of time in overall MVPA across weekend days. Following consultation with the lead author of the English child version of the YAP (also a co-author of this study-SF) a decision was made to ask the YAP-weekend item along with an overall weekday item, using the same wording (Table 1). The YAP was also used to also estimate sedentary behaviours whilst watching television, playing video games, using a mobile phone, a computer/tablet during COVID-19 restrictions. An additional question of ‘*doing school work*’ was also included to capture the amount of time children spent doing sedentary school work during COVID-19 restrictions (Table 1). A binary variable of meeting screen time (ST) guidelines (< 2hours a day)^34, 35^ was calculated by the values of each answer for sedentary screen behaviours (Table 1). Childrens average sleep time was estimated by children reporting their normal bedtime and a wake time. Sleep time was categorised into meeting sleep guidelines^34^ (9-to-11 hours a day) or more or less.

**Table 1:** Questionnaire items and processing methods used for during COVID-19 first lockdown analysis.

|  |  |  |  |  |
| --- | --- | --- | --- | --- |
| **Variable(s)** | **Question(s)** | **Response option** | **Processing variable** | **Source** |
| Sufficiently physically active (normally doing 60 minutes of moderate-vigorous physical activity everyday). | a) for a normal weekday (Monday, Tuesday, Wednesday, Thursday, Friday), during the last 7 days, how much physical activity did you do? (e.g. dancing, online exercise, games/sports, jobs at home, cycling).This can be anything that made you feel warmer, breathe harder or your heart beat faster.'<br>b) for a normal weekend day (Saturday, Sunday), during the last 7 days, how much physical activity did you do? (e.g. dancing, online exercise, games/sports, jobs at home, cycling).This can be anything that made you feel warmer, breathe harder or your heart beat faster.' | a) No activity (0min)<br>b) small amount activity (1 to 30 min)<br>c) small to moderate amount of activity (31 to 60 min)<br>d) moderate to large amount of activity (1 to 2 hours)<br>e) large amount of activity (more than 2 hours). | Children who selected either d) or e) (60 minutes or more) for both weekday and weekends were categorised as being sufficiently active (i.e. meeting guidelines of $\geq 60$ minutes of MVPA) or not. | Bespoke deriving of variable using Youth Activity Profile* |
| Sedentary - watching TV<br>Sedentary - playing video games<br>Sedentary - using computers/tablets<br>Sedentary - using mobile phone<br>Sedentary - doing school work | During the last 7 days, on a normal day, how many hours did you spend doing the following activities while sitting or lying down?<br>- watching TV (but not time spent play video games)<br>- playing video games (on a game console e.g. PlayStation or switch, mobile phone, tablet or computer)<br>- using computers/tablets (for social activity, e.g. social media, surfing web or video calling but not playing computer games or school work)<br>- using mobile phone (to talk, text or socialise, e.g. social media, but not playing games)<br>- doing school work (in books or on a computer/tablet (e.g. maths, reading, topic work) | a) No activity<br>b) 1 hour or less<br>c) 1-2 hours<br>d) 3-4 hours<br>e) >4 hours | Children were asked to report on a Likert scale (0 = no time, 5 = 4 hours or more) how much time they spent on a normal day in the last 7 days being sedentary (sitting, reclining or lying down) whilst watching television, playing video games, using a mobile phone, a computer/tablet. An additional question of '*doing school work*' was also included to capture the amount of time children spent doing sedentary school work during COVID-19 restrictions. Categories were collapsed into <1 hour, 1-3 hours, >3 hours, due to the small number of Responses for no activity and 4 hours or more. | Youth Activity Profile |
| Sedentary screen time | Variable made up of sedentary TV, video games, using computer/tablets, mobile phone<br>NOT school work | Derived variable | Each categorical was coded with a time amount (i.e. no time = 0, less than 1 hour = 0.5, 1-2 hours = 1.5, 3-4 hours = 3.5, 4 hours = 4). Summed all of the time amounts to estimate normal time spent using screen. Children with > 2 hours as not meeting screen time guidelines = 0, < 2 hours meeting ST-guidelines = 1. | Bespoke deriving of variable using Youth Activity Profile |
| Sleep | a) in the last seven days what time have you normally fallen asleep?<br>b) in the last seven days what time have you normally woken up? | Free text | Using the time from each answer a self-reported average sleep time was estimated for each child.<br>Time in hours was then coded into either not meeting - less than 9 hours, meeting - sleeping between 9-11 hours, not meeting - sleeping. | Bespoke |
| Frequency of leaving the home (including garden/yard) a day | *on a normal day in the last 7 days, how many times did you leave your home (away from your house and garden)?'* | a) Stayed at home<br>b) left home once a day<br>c) left the home more than once a day. | Used categories as asked | Bespoke |
| Duration of leaving the home (including garden/yard) a day | Children who reported leaving the home were asked - how long did you go for? | a) ≤30 minutes<br>b) 31-60 minutes<br>c) ≥60 minutes) | Used categories as asked | Bespoke |
| Usually did when leaving the home? | Children who reported leaving the home were asked - what did you usually do? (choose all that apply) | a)walk<br>b) run/jog<br>c) scoot/ride bike<br>d) other [free text] | Free text answers were analysed by four trained researchers who assessed and agreed upon the following additional categories: ; 'Sports and games and Other (non-active pursuits such as travelling to see family and friends) | Bespoke |
| Usually go when leaving the home? | Children who reported leaving the home were asked - where did you usually go? | a) Street<br>b) Park<br>c) Shops<br>d) other [free text] | Free text answers were analysed by four trained researchers who assessed and agreed upon the following additional categories: ; Greenspace/nature (e.g. woods, canals, moors, countryside) and Other neighbourhood areas (all other responses). | Bespoke |
| *Youth Activity Profile – English Child Version - Fairclough, S. J., Christian, D. L., Saint-Maurice, P. F., Hibbing, P. R., Noonan, R. J., Welk, G. J., Dixon, P. M., & Boddy, L. M. (2019). Calibration and Validation of the Youth Activity Profile as a Physical Activity and Sedentary Behaviour Surveillance Tool for English Youth. *International journal of environmental research and public health*, 16(19), 3711.<br>https://doi.org/10.3390/ijerph16193711 |  |  |  |  |

Because of the uniqueness of the COVID-19 lockdown and subsequent reduced opportunities for children to being physically active as would normally be, children were asked to answer questions on the frequency they normally left the home, the duration they would normally leave for, the type of activities they usually did when leaving the home, and where they usually went (Table 1).

#### Before COVID-19

For the sub-sample of children with available data from before COVID-19, PA levels were measured by children completing the PAQ-C with the support of trained researchers during school time (2017-2019). The PAQ-C is a validated PA seven day recall questionnaire, that measures general levels of MVPA of children aged 8-14 years by assessing participation in different physical activities as well as activity during physical education, lunch break, recess (play time), before school, after school, evenings and weekends.^33, 36^ The scoring of the PAQ-C is based upon an average of all questions asked, with a score between 1 (low activity rating) to 5 (high activity rating).^33, 36^ Cut-off values indicating whether children were sufficiently active (relating to cardio-respiratory fitness^37^) were applied (2.7 aggregate score [out of 5] for girls, 2.9 aggregate score [out of 5] for boys).

### Statistical analysis

Descriptive statistics for all variables were generated. Continuous variables were described using mean and standard deviation; categorical variables using counts and proportions. For aim one (whole sample, during COVID-19) univariate statistical tests were performed (Pearson Chi-square tests (χ^2^), with Bonferroni-adjusted *p-*values, independent *t*-tests, one-way analysis of variance and non-parametric alternatives) to examine whether there were differences between the outcome variable (sufficiently active [normally doing 60mins of MVPA a day]- yes or no) and independent variables (meeting sleep guidelines, time spent in sedentary behaviours, frequency and duration of leaving the home, and destination and type of active outside of the home. Because of the inequalities between sex and ethnic groups, univariate associations were examined between all measures with sex and ethnicity categories. Four multivariable logistic regressions were generated for the outcome (sufficiently active [normally doing 60mins of MVPA a day]- yes or no). The first model included key demographic variables (age, sex, ethnicity, IMD) and whether children still attended school. The second model added the five sedentary behaviours to demographics variables. The third model included the frequency children reported of leaving the home. The fourth and final model included only children who reported leaving the home and included the variable of duration of time away from the home, destination children usually went to, and type of PA children did when away from the home. For aim two change over time from baseline (pre-COVID-19) to follow-up (during COVID-19) for children being sufficiently active (binary 0 for ‘No’ and 1 for ‘Yes’) was investigated using the McNemar test for significance of changes on the subsample of children with data available at the time at different time points (pre-COVID-19, during the first COVID-19 lockdown). Potential demographic factors, associated with any significant change in compliance (sex, age difference [months] between pre-post COVID, ethnicity, IMD) were investigated using logistic regression through simultaneous entry of independent variables. The outcome variable was coded 0 for the ‘absence of negative change’ for being sufficiently active and 1 for the ‘presence of negative change’ being sufficiently active. All analysis was conducted using Stata v15.0 (StataCorp., College Station, TX).

## Results

### Descriptive Statistics

Out of a total n=5,298 eligible children, n=970 (18.3%) of children agreed to take part, completed and returned a survey in spring 2020. A total of n=949 (17.9%) had completed PA data and were included in the analysis for aim 1. A total of n=634 (66.8%, based upon 949 children) children had matched PA data prior to COVID-19. The characteristics for both pre and during COVID-19 samples are reported in Table 2.

**Table 2:** Study samples demographics and characteristics.

| **Table 2: Study samples demographics and characteristics.** |  |  |
| --- | --- | --- |
|  | During COVID-19<br>sample<br>(n=949) | Pre-COVID-19<br>sample<br>(n=643) |
| **Age, m (SD)\*** | 10.5 (1.1) | 9.1 (1.1) |
| **Sex, n (%)** |  |  |
| Male | 486 (51.2) | 321(50.6) |
| Female | 463 (48.8) | 313 (49.4) |
| **Ethnicity, n (%)** |  |  |
| White British | 385 (40.6) | 254 (40.1) |
| Pakistani Heritage | 418 (44.1) | 275 (43.4) |
| Other Ethnicity | 146 (15.4) | 105 (16.6) |
| **Index of Multiple Deprivation, n (%)** |  |  |
| Most deprived nationally | 355 (37.4) | 237 (37.4) |
| 2nd most deprived nationally | 140 (14.8) | 97 (15.3) |
| 3rd most deprived nationally | 166 (17.5) | 114 (18.0) |
| 4th < most deprived nationally | 288 (30.5) | 186 (29.3) |
| **Attending School, n (%)** |  |  |
| Yes | 95 (10.0) | 67 (1.6) |
| No | 854 (90.0) | 563 (89.4) |
| The average age between pre and during COVID-19 first lockdown was 1.2 years (0.72) /15.2 months (8.6) |  |  |

### During COVID-19 lockdown: Levels of self-reported physical activity and activity away from the home

Twenty-seven per cent of children reported being sufficiently active (>60 min MVPA daily) during the first COVID-19 lockdown (Table 3). Children reported spending an average of 10.6 hours (SD=1.5) a day sleeping, with 69% meeting sleep guidelines of 9 to 11 hours/day. Almost one third of children reported spending ≥3 hours a day doing sedentary schoolwork (32.9%) and more than ≥3 hours a day playing sedentary video games (29.6%), and a majority of children did not meet screen time guidelines (89.9%). The majority of children reported that they had usually left the home environment during the previous seven days, with 53.9% leaving once a day, and 16.7% more than once a day. However, 30% reported that they had normally stayed at home. Of the children who reported leaving the home at least once a day, the majority of children reported leaving between 31-60 minutes (54%). The most frequently reported type of activities outside of the home was walking (77%) and riding a bike/scooter (41.9%), and the most frequent reported places for children to go was the street (33.7%) and park (34.5%).

**Table 3:** Levels, sex and ethnic differences of childrens self-reported physical activity, usual sleep duration, sedentary behaviours, whether attending school, frequency and duration of leaving the home environment during a COVID-19 UK restrictions (April-June 2020).

|  | All (n=946) | Males (n=486) | Females (n=463) | *p* | White British (n=385) | Pakistani Heritage (n=418) | Other (n=146) | *p* |
| --- | --- | --- | --- | --- | --- | --- | --- | --- |
| Physical activity self-reported during COVID-19 first lockdown (MVPA - ≥ 60 minutes daily), n (%) |  |  |  |  |  |  |  |  |
| Yes - sufficiently active | 259 (27.4) | 145 (29.8) | 114 (24.8) | .08 | 131 (34.1) | 95 (22.8) | 33 (22.8) | .00 |
| Not - sufficiently active | 687 (72.6) | 341 (70.2) | 246 (75.2) |  | 253 (65.9) | 322 (77.2) | 112 (77.2) |  |
| Attending School, n (%) |  |  |  |  |  |  |  |  |
| Yes | 95 (10.0) | 56 (11.5) | 39 (8.4) | .11 | 54 (14.0) | 25 (6.0) | 16 (10.7) | .00 |
| No | 854 (90.0) | 430 (88.5) | 424 (91.6) |  | 331 (86.0) | 393 (94.02) | 130 (89.0) |  |
| Meeting Sleep guidelines, n (%) |  |  |  |  |  |  |  |  |
| Not meeting - less than 9 hours | 63 (6.8) | 39 (8.3) | 24 (5.3) | .00 | 29 (7.7) | 22 (5.5) | 12 (8.5) | .00 |
| Yes- meeting guidelines - 9-11 hours | 637 (68.9) | 336 (71.3) | 301 (66.5) |  | 303 (80.2) | 239 (59.1) | 95 (66.9) |  |
| Not meeting - Sleep more than 11 hours | 224 (24.2) | 96 (20.4) | 128 (28.3) |  | 46 (12.1) | 143 (35.4) | 35 (24.6) |  |
| Sedentary- Watching Television (not time playing video games), n (%) |  |  |  |  |  |  |  |  |
| < 1 hour | 393 (42.3) | 211 (44.4) | 182 (40.0) | .26 | 160 (42.7) | 173 (41.9) | 60 (42.3) | .46 |
| 1-3 hours | 380 (40.9) | 182 (38.3) | 198 (43.5) |  | 160 (42.7) | 160 (38.7) | 60 (42.3) |  |
| 3hr < | 157 (16.9) | 82 (17.3) | 75 (16.5) |  | 55 (14.6) | 80 (19.4) | 22 (15.4) |  |
| Sedentary - Video games on a games console, n (%) |  |  |  |  |  |  |  |  |
| < 1 hour | 365 (38.9) | 124 (25.8) | 241 (52.6) | .00 | 117 (30.7) | 176 (42.7) | 72 (49.7) | .00 |
| 1-3 hours | 295 (31.5) | 157 (32.7) | 138 (30.1) |  | 120 (31.5) | 137 (33.3) | 38 (26.2) |  |
| 3hr < | 278 (29.6) | 199 (41.5) | 79 (17.3) |  | 144 (37.8) | 99 (24.0) | 35 (24.1) |  |
| Sedentary - Computers/tablets use for social activity, n (%) |  |  |  |  |  |  |  |  |
| < 1 hour | 653 (70.6) | 338 (71.9) | 315 (69.23) | .56 | 259 (69.4) | 283 (69.0) | 111 (78.2) | .309 |
| 1-3 hours | 176 (19.03) | 83 (17.7) | 93 (20.4) |  | 74 (19.8) | 81 (19.8) | 21 (14.8) |  |
| 3hr < | 96 (10.38) | 49 (10.4) | 47 (10.3) |  | 40 (10.7) | 46 (11.2) | 10 (7.0) |  |
| Sedentary - Mobile phone use (not playing games), n (%) |  |  |  |  |  |  |  |  |
| < 1 hour | 707 (76.4) | 386 (81.4) | 321 (71.0) | .00 | 224 (64.9) | 342 (83.4) | 121 (86.4) | .00 |
| 1-3 hours | 123 (13.3) | 55 (11.6) | 68 (15.0) |  | 78 (20.7) | 32 (7.8) | 13 (9.3) |  |
| 3hr < | 96 (10.4) | 33 (7.0) | 63 (13.9) |  | 54 (14.4) | 36 (8.8) | 6 (4.3) |  |
| Sedentary - School Work (books, computers), n (%) |  |  |  |  |  |  |  |  |
| < 1 hour | 271 (28.8) | 143 (29.7) | 128 (27.9) | .01* | 100 (26.3) | 134 (32.4) | 37 (25.3) | .00 |
| 1-3 hours | 360 (38.3) | 201 (41.8) | 159 (34.6) |  | 128 (33.7) | 170 (41.1) | 62 (42.5) |  |
| 3hr < | 309 (32.87) | 137 (28.5) | 172 (37.5) |  | 152 (40.0) | 110 (26.6) | 47 (32.2) |  |
| Screen time - Meeting Guidelines ≤2 Hours |  |  |  |  |  |  |  |  |
| Not Meeting | 842 (89.9) | 438 (91.4) | 404 (88.2) | .10 | 353 (93.6) | 364 (87.3) | 125 | .00 |
|  |  |  |  |  |  |  | (87.4) |  |
| Meeting | 95 (10.1) | 41 (8.6) | 54 (11.8) |  | 24 (6.4) | 53 (12.7) | 18 (12.6) |  |
| Frequency of leaving home (including garden/yard) a day, n (%) |  |  |  |  |  |  |  |  |
| Stayed at home | 279 (29.7) | 145 (30.0) | 134 (29.4) | .10 | 68 (17.9) | 163 (39.5) | 48 (32.9) | .00 |
| Once a day | 507 (53.9) | 260 (53.7) | 247 (54.2) |  | 244 (64.0) | 193 (46.7) | 70 (48.0) |  |
| More than once a day | 154 (16.4) | 79 (16.3) | 75 (16.5) |  | 69 (18.1) | 57 (13.8) | 28 (19.2) |  |
| Duration of time away from home (including garden/yard) a day, n(%) |  |  |  |  |  |  |  |  |
| < 30 minutes | 100 (15.0) | 44 (12.9) | 56 (17.2) | .17 | 30 (9.6) | 56 (22.1) | 14 (14.3) | .00 |
| 31-60 minutes | 359 (54.0) | 182 (53.5) | 177 (54.5) |  | 163 (51.9) | 141 (55.7) | 55 (56.1) |  |
| 60 minutes < | 206 (31.0) | 114 (33.5) | 92 (28.3) |  | 121 (38.5) | 56 (22.1) | 29 (29.6) |  |
| Where did children usually go when leaving the home? n(%) |  |  |  |  |  |  |  |  |
| Street | 221 (33.7) | 106 (31.2) | 115 (36.4) | .37 | 101 (32.6) | 91 (36.7) | 29 (29.9) | .00 |
| Shops | 64 (9.8) | 31 (9.1) | 33 (10.4) |  | 22 (7.1) | 31 (12.5) | 11 (11.3) |  |
| Park | 226 (34.5) | 118 (34.8) | 108 (34.2) |  | 83 (26.8) | 103 (41.5) | 40 (41.2) |  |
| Greenspace/nature (e.g. woods, local fields) | 80 (12.2) | 36 (10.6) | 28 (8.9) |  | 67 (21.6) | 6 (2.4) | 7 (7.2) |  |
| Other neighbourhood areas | 64 (9.8) | 48 (14.2) | 32 (10.1) |  | 37 (11.9) | 17 (6.9) | 10 (10.3) |  |
| What did children usually do when leaving home? - Walk n(%) |  |  |  |  |  |  |  |  |
| No – did not walk | 154 (23.1) | 93 (27.1) | 61 (18.9) | .01 | 54 (17.1) | 75 (29.5) | 25 (25.3) | .00 |
| Yes – did walk | 514 (77.0) | 250 (72.9) | 264 (81.2) |  | 261 (82.9) | 179 (70.5) | 74 (74.8) |  |
| What did children usually do when leaving home? – Run/Jog n(%) |  |  |  |  |  |  |  |  |
| No – did not Run/Jog | 535 (80.1) | 263 (76.7) | 272 (83.7) | .02* | 259 (82.2) | 207 (81.5) | 69 (69.7) | .02* |
| Yes – did Run/Jog | 133 (19.9) | 80 (23.3) | 53 (16.3) |  | 56 (17.8) | 47 (18.5) | 30 (30.3) |  |
| What did children usually do when leaving home? – Ride bike/scoot n(%) |  |  |  |  |  |  |  |  |
| No – did not Ride bike/scoot | 388 (58.1) | 187 (54.5) | 201 (61.9) | .06 | 171(54.3) | 164 (64.6) | 53 (53.5) | .03* |
| Yes – did Ride bike/scoot | 280 (41.9) | 156 (45.5) | 124 (38.1) |  | 144 (45.7) | 90 (35.4) | 46 (46.5) |  |
| What did children usually do when leaving home? –Play, Sports or Games (e.g. playing n(%) |  |  |  |  |  |  |  |  |
| No – did play sports or games | 612 (91.6) | 308 (89.8) | 304 (93.5) | .08 | 296 (94.0) | 230 (90.6) | 86 (86.9) | .06 |
| Yes – did play sports or games | 56 (8.4) | 35 (10.2) | 21 (6.5) |  | 19 (33.9) | 24 (9.5) | 13 (13.1) |  |
| What did children usually do when leaving home? – Other (e.g. travelling in car) n(%) |  |  |  |  |  |  |  |  |
| No – did not Ride bike/scoot | 636 (95.2) | 330 (96.2) | 306 (94.2) | .21 | 305 (96.8) | 238 (93.7) | 93 (93.9) | .18 |
| Yes – did Ride bike/scoot | 32 (4.8) | 13 (3.8) | 19 (5.9) |  | 10 (3.2) | 16 (6.3) | 6 (6.1) |  |
| * Non-significant due to Bonferroni correction |  |  |  |  |  |  |  |  |

### During COVID-19 lockdown: Sex and ethnicity behaviour differences during the COVID-19 lockdown

Univariate sex and ethnicity differences are reported in Table 3. Differences between boys and girls were found for sleep duration (≥11 hours: Girls=28.3% > Boys=20.4%); time spent normally playing console video games (≥3 hours: Boys=41.5% > Girls=17.3); using mobile phones (≥3 hours: Girls=13.9% > Boys=7.0); and usually walking (type of activity) when outside of the home (Girls=81.2% > Boys=72.9%). Differences between ethnic groups were found for being sufficiently active (WB=34.1% vs P=22.8% vs O=22.8%), still attending school (WB=14% vs. P=10.7% and O=6%); sleep duration (9-11 hours: WB=80.2% vs PH=59.1% vs O=66.9); time spent normally - playing console video games (≥3 hours :WB=37.8% vs PH=24.% vs O=24.1%), using mobile phones (≥3 hours: WB=35.1% vs PH=16.6% vs O=13.6%), meeting ST-guidelines (<2hours WB=6.4% vs PH=12.7% vs 12.6%); frequency of leaving the home (stayed at home: PH=39.5% vs O=32.9% vs WB=17.9%); duration of time leaving the home (≥60 minutes: WB=38.5% vs PH=22.1% vs O=29.6%); places children usually went outside of the home (Park: PH and O =41.5% vs WB=26.8%, Greenspace/nature: WB=21.6% vs PH=2.4% vs O=7.2%); and usually walking when outside of the home (WB=82.9% vs PH=70.5% vs O=74.8%).

### During COVID-19 lockdown: Factors associated with children being sufficiently active during COVID-19 lockdown

Univariate factors between children’s self-reported PA and predictor variables (Table 4) were age, ethnicity, duration of playing video games on a console, normal daily frequency of leaving the home, normal daily duration of leaving the home, the place children usually went to outside of the home, and if they took part in running/jogging, riding a bike/scooter, and playing sports and games.

**Table 4:** Univariate analysis of difference between children sufficiently physically active (< 60 minutes usually a day) with demographics, and independent variables during COVID-19 UK restrictions.

|  |  |  |  |
| --- | --- | --- | --- |
|  | Sufficiently physically active |  |  |
|  | Yes n=259 (27.4% ) | No n=687 (72.6%) | p |
| Age, m (SD) | 10.3 (1.1) | 10.6 (1.1) | .01 |
| Gender, n (%) |  |  |  |
| Male | 145 (29.8) | 341 (70.2) | .08 |
| Female | 114 (24.8) | 246 (75.2) |  |
| Ethnicity, n (%) |  |  |  |
| White British | 131 (34.1) | 253 (65.9) | .00 |
| Pakistani Heritage | 95 (22.8) | 322 (77.2) |  |
| Other ethnicities | 33 (22.8) | 112 (77.2) |  |
| Index of Multiple Deprivation, n (%) |  |  |  |
| Most deprived nationally | 81 (22.9) | 273 (77.1) | .02* |
| 2nd most deprived nationally | 43 (30.7) | 97 (69.3) |  |
| 3rd most deprived nationally | 40 (24.2) | 125 (75.8) |  |
| 4th < most deprived nationally | 95 (33.1) | 192 (66.9) |  |
| Attending School, n (%) |  |  |  |
| Yes | 34 (35.8) | 61 (64.21) | .05 |
| No | 225 (26.4) | 626 (73.7) |  |
| Meeting Sleep guidelines - self reported, n (%) |  |  |  |
| Not meeting - less than 9 hours | 16 (25.4) | 47 (74.6) | .01** |
| Yes- meeting guidelines - 9-11 hours | 193 (30.4) | 443 (69.7) |  |
| Sleep more than 11 hours | 45 (20.1) | 170 (80.9) |  |
| Watching Television (not time playing video games), n (%) |  |  |  |
| < 1 hour | 112 (28.6) | 219 (71.4) | .040*** |
| 1-3 hours | 111 (29.3) | 268 (70.7) |  |
| 3hr < | 30 (19.1) | 127 (80.9) |  |
| Video games on a games console, n (%) |  |  |  |
| < 1 hour | 109 (30.0) | 155 (70.0) | .00 |
| 1-3 hours | 95 (32.4) | 198 (67.6) |  |
| 3hr < | 53 (19.1) | 225 (80.9) |  |
| Computers/tablets use for social activity, n (%) |  |  |  |
| < 1 hour | 189 (28.9) | 466 (71.2) | .12 |
| 1-3 hours | 49 (27.8) | 127 (72.2) |  |
| 3hr < | 18 (28.7) | 79 (81.4) |  |
| Mobile phone use (not playing games), n (%) |  |  |  |
| < 1 hour | 199 (28.2) | 506 (71.8) | .14 |
| 1-3 hours | 36 (29.3) | 87 (13.0) |  |
| 3hr < | 18 (19.0) | 77 (11.5) |  |
| School Work (books, computers), n (%) |  |  |  |
| < 1 hour | 69 (25.3) | 204 (74.7) | .10 |
| 1-3 hours | 90 (25.0) | 270 (75.0) |  |
| 3hr < | 100 (32.3) | 210 (67.7) |  |
| Screen time - Meeting Guidelines ≤2 Hours |  |  |  |
| Not Meeting | 72 (10.6) | 607 (89.4) | .38 |
| Meeting | 232 (8.7) | 232 (91.3) |  |
| Frequency of leaving home (including garden/yard) a day, n (%) |  |  |  |
| Stayed at home | 46 (16.6) | 232 (83.5) | .00 |
| Once a day | 149 (29.4) | 358 (70.6) |  |
| More than once a day | 62 (40.3) | 92 (59.7) |  |
| Duration of time away from home (including garden/yard) a day, n(%) |  |  |  |
| < 30 minutes | 9 (9.0) | 91 (91.0) | .00 |
| 31-60 minutes | 91 (25.4) | 268 (74.7) |  |
| 60 minutes < | 111 (53.9) | 95 (46.1) |  |
| Where did children usually go when leaving the home? n(%) |  |  |  |
| Street | 66 (29.9) | 155 (70.1) | .00 |
| Shops | 6 (9.4) | 58 (90.6) |  |
| Park | 77 (34.2) | 148 (65.8) |  |
| Greenspace/nature (e.g. woods, local fields) | 34 (42.5) | 46 (57.5) |  |
| Other neighbourhood areas | 27 (42.2) | 37 (57.8) |  |
| What did children usually do when leaving home? - Walk n(%) |  |  |  |
| No – did not walk | 47 (30.5) | 107 (69.5) | .70 |
| Yes – did walk | 165 (32.2) | 348 (67.8) |  |
| What did children usually do when leaving home? – Run/Jog n(%) |  |  |  |
| No – did not Run/Jog | 155 (29.0) | 379 (70.1) | .00 |
| Yes – did Run/Jog | 57 (42.9) | 76 (57.1) |  |
| What did children usually do when leaving home? – Ride bike/scoot n(%) |  |  |  |
| No – did not Ride bike/scoot | 99 (25.8) | 288 (74.4) | .00 |
| Yes – did Ride bike/scoot | 113 (40.4) | 167 (59.6) |  |
| What did children usually do when leaving home? –Play, Sports or Games (e.g. playing n(%) |  |  |  |
| No – did play sports or games | 186 (30.4) | 425 (69.6) | .01 |
| Yes – did play sports or games | 26 (46.4) | 30 (53.6) |  |
| What did children usually do when leaving home? – Other things (e.g. travel in car) n(%) |  |  |  |
| No | 205 (32.3) | 430 (67.7) | .01 |
| Yes | 7 (21.9) | 25 (78.1) |  |
| * corrected p-value = .006, non-significant |  |  |  |
| ** corrected p-value = .008, non-significant |  |  |  |
| *** corrected p-value = .008, non-significant |  |  |  |

For the multivariable analysis, summaries of logistic regression models (1, 2, 3, 4) are reported in Table 5 (a full results table is found in Appendix 1 – supplementary material). In model 1, variables that decreased the odds of being sufficiently active were age (years) (OR=0.82, 95%CI 0.72-0.94), and ethnicity (reference: WB); PH children (OR 0.64, 95%CI 0.44-0.92), Other (OR=0.57, 95%CI 0.35-0.90). In model 2 (which included sedentary behaviours), being a girl (OR=0.63, 95%CI 0.45-0.88) and playing on video games ≥3 hours a day (OR=0.43, 95%CI 0.28-0.67) significantly decreased the odds of being sufficiently active, in addition to age and ethnicity. In model 3 (which included daily frequency of leaving the home), age, being a girl, being from another ethnic group and playing video games (≥3 hours a day) still decreased the odds of being sufficiently active; however, being of PH no longer did. Leaving the home at least once a day significantly increased the odds (OR=1.57 95%CI(1.04-2.36)), with the odds increasing further for children who reported leaving the home more than once a day (OR=2.73, 95%CI 1.66-4.48). In model 4, (which included duration, place and type of activity), age and playing videos for ≥3 hours/day significantly decreased the odds, but leaving the home for 31-60 minutes significantly increased the odds (OR=2.21, 95%CI 1.01-4.8), and the odds increased further for children reporting leaving the home for ≥60 minutes (OR=7.9, 95%CI 3.5-18.0). Children reporting that the place they usually went too was the shop which reduced the odds of children being sufficiently active (OR=0.36, 95%CI 0.13-0.98). Odds were increased for children reporting that usually took part in running/jogging (OR=2.13, 95%CI 1.30-3.47), riding a bike/scooter (OR=1.52, 95%CI 1.01-2.31), and playing sports and games (OR=2.13, 95%CI 3.4-2.70).

**Table 5:** Multivariable logistic regression analysis of factors (demographic, self-reported sleep duration and sedentary behaviours, school attendance, frequency, duration, type of activity and place of destination of children when leaving the home environment), with children self-reporting being sufficiently physically active (>60 minutes usually a day) during COVID-19 UK restrictions (April-June 2020).

|  | Model 1 (n=946) |  |  | Model 2 (n=875) |  |  | Model 3 (n=868) |  |  | Model 4 (n=602) |  |  |
| --- | --- | --- | --- | --- | --- | --- | --- | --- | --- | --- | --- | --- |
|  | OR | (95% CI) | *p* | OR | 95% CI | *p* | OR | 95% CI | *p* | OR | 95% CI | *p* |
| **Age (years)** | 0.82 | (0.72-0.94) | **0.00** | 0.83 | (0.72-0.97) | **0.02** | 0.83 | (0.12-0.97) | **0.02** | 0.81 | (0.67-0.99) | **0.04** |
| **Sex- Male (Reference)** |  |  |  |  |  |  |  |  |  |  |  |  |
| Female | 0.81 | 0.60-1.08 | 0.15 | 0.63 | (0.45-0.88) | **0.01** | 0.61 | (0.44-0.86) | **0.01** | 0.76 | (0.50-1.17) | 0.22 |
| **Ethnicity - White British (Reference)** |  |  |  |  |  |  |  |  |  |  |  |  |
| Pakistani Heritage | 0.64 | (0.44-0.92) | **0.02** | 0.62 | (0.41-0.95) | **0.03** | 0.72 | (0.47-1.12) | 0.14 | 0.82 | (0.46-1.48) | 0.51 |
| Other | 0.57 | (0.35-0.90) | **0.02** | 0.50 | (0.30-0.83) | **0.01** | 0.53 | (0.31-0.90) | **0.02** | 0.48 | (0.24-0.94) | **0.03** |
| **Index of Multiple Deprivation Most Deprived (Reference)** |  |  |  |  |  |  |  |  |  |  |  |  |
| 2nd most deprived | 1.46 | (0.94-2.28) | 0.09 | 1.30 | (0.81-2.08) | 0.28 | 1.22 | (0.76-1.98) | 0.41 | 1.41 | (0.78-2.54) | 0.26 |
| 3rd most deprived | 0.97 | (0.62-1.52) | 0.91 | 0.78 | (0.48-1.26) | 0.31 | 0.82 | (0.51-1.33) | 0.43 | 0.84 | (0.45-1.57) | 0.58 |
| 4th < most deprived | 1.33 | (0.89-2.00) | 0.17 | 1.13 | (0.72-1.75) | 0.60 | 1.12 | (0.71-1.75) | 0.63 | 1.40 | (0.81-2.41) | 0.23 |
| **Attending School - Yes (Reference)** |  |  |  |  |  |  |  |  |  |  |  |  |
| No | 1.41 | (0.89-2.23) | 0.14 | 1.32 | (0.79-2.18) | 0.29 | 1.16 | (0.69-1.96) | 0.57 | 1.33 | (0.72-2.47) | 0.36 |
| **Meeting Sleep guidelines Not meeting (Reference)** |  |  |  |  |  |  |  |  |  |  |  |  |
| Yes- meeting (9-11 hr) |  |  |  | 1.32 | (0.69-2.54) | 0.40 | 1.31 | (0.67-2.55) | 0.43 | 1.59 | (0.69-3.73) | 0.29 |
| 11 hours < |  |  |  | 0.87 | (0.42-1.81) | 0.72 | 0.88 | (0.42-1.13) | 0.74 | 1.43 | (0.55-3.70) | 0.47 |
| **Watching Television < 1 hour (Reference)** |  |  |  |  |  |  |  |  |  |  |  |  |
| 1-3 hours |  |  |  | 1.04 | (0.75-1.46) | 0.80 | 1.08 | (0.77-1.52) | 0.65 | 1.17 | (0.76-1.79) | 0.48 |
| 3hr < |  |  |  | 0.74 | (0.46-1.19) | 0.21 | 0.69 | (0.43-1.23) | 0.14 | 0.74 | (0.41-1.33) | 0.31 |
| **Video games on a games console** |  |  |  |  |  |  |  |  |  |  |  |  |
| < 1 hour (Reference) |  |  |  |  |  |  |  |  |  |  |  |  |
| 1-3 hours |  |  |  | 0.91 | (0.63-1.32) | 0.62 | 0.92 | (0.63-1.33) | 0.67 | 0.94 | (0.58-1.51) | 0.95 |
| 3hr < |  |  |  | 0.43 | (0.28-0.67) | 0.00 | 0.45 | (0.29-0.70) | 0.00 | 0.52 | (0.30-0.90) | 0.02 |
| **Computers/tablets use for social activity**<br>< 1 hour (Reference) |  |  |  |  |  |  |  |  |  |  |  |  |
| 1-3 hours |  |  |  | 1.11 | (0.74-1.68) | 0.61 | 1.08 | (0.71-1.63) | 0.72 | 1.34 | (0.81-2.33) | 0.25 |
| 3hr < |  |  |  | 0.43 | (0.44-1.45) | 0.46 | 0.86 | (0.47-0.71) | 0.64 | 1.67 | (0.72-3.86) | 0.45 |
| **Mobile phone use**<br>< 1 hour (Reference) |  |  |  |  |  |  |  |  |  |  |  |  |
| 1-3 hours |  |  |  | 1.03 | (0.64-1.66) | 0.90 | 1.11 | (0.69-1.79) | 0.68 | 1.04 | (0.58-1.89) | 0.88 |
| 3hr < |  |  |  | 0.73 | (0.39-1.37) | 0.33 | 0.81 | (0.43-1.52) | 0.51 | 0.73 | (0.31-1.68) | 0.45 |
| **School Work**<br>< 1 hour (Reference) |  |  |  |  |  |  |  |  |  |  |  |  |
| 1-3 hours |  |  |  | 0.89 | (0.60-1.32) | 0.56 | 0.89 | (0.59-1.32) | 0.56 | 0.85 | (0.51-1.41) | 0.52 |
| 3hr < |  |  |  | 1.20 | (0.80-1.80) | 0.37 | 1.20 | (0.79-1.81)) | 0.39 | 1.03 | (0.61-1.73) | 0.91 |
| **Frequency of leaving home**<br>Stayed at Home (Reference) |  |  |  |  |  |  |  |  |  |  |  |  |
| Once a day (Reference - Model 4) |  |  |  |  |  |  | 1.57 | (1.04-2.36) | 0.03 |  |  |  |
| More than once a day |  |  |  |  |  |  | 2.73 | (1.66-4.48) | 0.00 | 1.08 | (0.67-1.73) | 0.75 |
| **Duration away from home**<br>< 30 minutes (Reference) |  |  |  |  |  |  |  |  |  |  |  |  |
| 31-60 minutes |  |  |  |  |  |  |  |  |  | 2.21 | (1.01-4.8) | 0.04 |
| 60 minutes < |  |  |  |  |  |  |  |  |  | 7.9 | (3.5-18.0) | 0.00 |
| **Where did children usually go when leaving the home?**<br>Street (reference) |  |  |  |  |  |  |  |  |  |  |  |  |
| Shops |  |  |  |  |  |  |  |  |  | 0.36 | (0.13-0.98) | 0.04 |
| Park |  |  |  |  |  |  |  |  |  | 1.09 | (0.68-1.77) | 0.72 |
| Greenspace/nature |  |  |  |  |  |  |  |  |  | 0.97 | (0.49-1.88) | 0.92 |
| Other neighbourhood areas |  |  |  |  |  |  |  |  |  | 0.90 | (44.3-1.85) | 0.78 |
| Usually do, Walk - No(reference) |  |  |  |  |  |  |  |  |  | 1.15 | (0.70-1.88) | 0.59 |
| Yes |  |  |  |  |  |  |  |  |  |  |  |  |
| **Usually do, Run/Jog -**<br>No (reference)<br>Yes |  |  |  |  |  |  |  |  |  | 2.13 | (1.30-3.47) | **0.00** |
| **Usually do, Ride bike/scoot -**<br>No (reference)<br>Yes |  |  |  |  |  |  |  |  |  | 1.52 | (1.01-2.31) | **0.04** |
| **Usually do, Play, Sports or Games**<br>- No (reference)<br>Yes |  |  |  |  |  |  |  |  |  | 2.13 | (1.1-4.31) | **0.03** |
| **Usually do, Other -** No (reference)<br>Yes) |  |  |  |  |  |  |  |  |  | 0.96 | (0.34-2.70) | 0.93 |
| Constant | 3.67 | (0.86-15.64) | 0.078 | 4.39 | (0.78-24.83) | 0.09 | 2.64 | (0.44-15.85) | 0.29 | 0.6748692 | (0.06-7.25) | 0.75 |
| Log likelihood | -539.2033 |  |  | -483.34 |  |  | -471.39388 |  |  | -310.63993 |  |  |
| Pseudo r-square | 0.029 |  |  | 0.062 |  |  | 0.077 |  |  | 0.1758 |  |  |
| Likelihood-Ratio chi-square (df) | 32.17 (8), p=0.000 |  |  | 63.34 (20), p=.0001 |  |  | 78.85(22), p=.0000 |  |  | 106.86(32), p=0.000 |  |  |

### Changes in children being sufficiently physically active before and during the COVID-19 lockdown

The sub-samples’ pre-COVID-19 PA (sub-sample, n=643) was PAQ-C score 3.2 (SD=0.77), with 69.4% (n=440) found to be sufficiently active. During COVID-19 the proportion of children being sufficiently active reduced to 28.7% (n=183). The magnitude of change was statistically significant (see Table 6), with 47.5% of children changing from being sufficiently active before COVID-19 to not being sufficiently active during COVID-19. A small number of children who were not sufficiently active pre-COVID did report being sufficiently during COVID-19 (7.0%, n=44), leading to a 40.5% reduction. A logistic regression model (Table 3) predicted that the age difference between the two measurement periods, ethnicity and sex did not significantly increase or decrease the odds of children negatively changing from being sufficiently active from before COVID-19 to during COVID-19.

**Table 6:** McNemar test for significance of changes in reported physical activity before COVID-19 and during COVID-19.

|  |  |  |  |  |  |
| --- | --- | --- | --- | --- | --- |
| Baseline n(%) | Follow-Up n(%) |  | McNemar test statistic |  |  |
|  | Sufficiently active | Not sufficiently active | X<sup>2</sup> | df | P |
| Sufficiently active | 139 (21.8%) | 301 (47.5%) | 191.5 | 1 | 0.00* |
| Not sufficiently active | 44 (7%) | 150 (23.7%) |  |  |  |

**Table 7:**
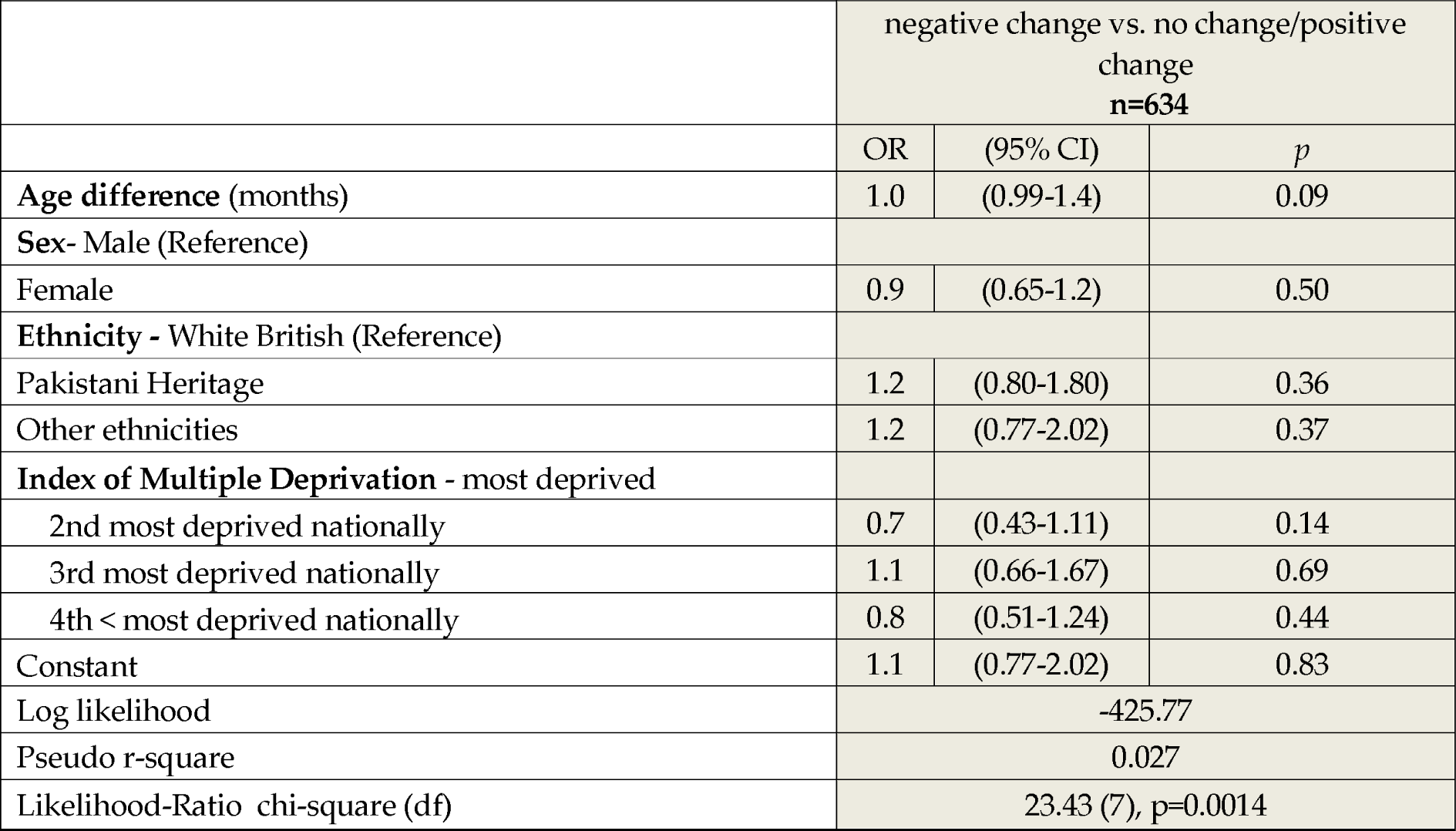
Factors for change in children being sufficiently physically active measured by self-report before and during COVID-19.

## Discussion

The purpose of this study was to investigate the levels, factors associated and change of childrens self-reported PA during the first COVID-19 in England. Results show levels of children reporting being sufficiently active has drastically reduced from before COVID-19. Factors associated with meeting guidelines during the first COVID-19 lockdown were child’s age, ethnicity (Pakistani Heritage and Other ethnic minorities [-]), sex (girls), self-reported video game usage (>3 hours a day[-]), and the frequency (>1 a day[+]), duration (>31 minutes[+]), type of activity (run/jog, ride bike/scooter, play, sports or games [+]) and place visited when leaving the home environment (shops [-]).

Only a quarter of children reported being sufficiently active enough to benefit their health during the first COVID-19 lockdown, and this reduced greatly from before COVID-19, independently of increased age. These findings are similar to other studies ^19, 21, 38^ and are unsurprising when considering the sharp change in the systems in which children’s PA would usually occur (i.e. school, sport clubs, parks, playgrounds, active travel). Daily PA outside of the home environment was allowed and has been consistently allowed by the UK-government during the first lockdown and throughout the pandemic, but not actively promoted.^38, 39^ This is unsurprising due to the priority being to reduce mixing of individual households. As the BiB COVID-19 study^26^ progresses further studies will be able to report on changes in PA during the pandemic and during differing restriction circumstances. It is likely, given the ongoing restrictions, that PA levels will remain lower than pre-pandemic. The short- and long-term health implications for reduced PA for a sustained period of time during childhood is unknown and this is something which the cohort study aims to investigate. Early life is particularly important for habit formation and has been shown that PA tracks from across the life course of young people^40–42^ so there is a possibility of long term health implications associated with reduced PA across the lifespan, triggered by reduced PA during the COVID-19 pandemic. This is something that requires careful monitoring and preventative interventions to reduce the likelihood of ongoing low PA levels.

Previous non-pandemic research has shown an association between children’s PA levels and time spent outdoors,^43, 44^ the current studies findings highlight how important time away from the home environment was for being active. Worryingly, 29.7% of children reported that they didn’t leave the home on a usual day during lockdown and this was strongly associated with not being sufficiently active (OR=1.6 once a day, OR=2.7 ≥once a day). For those who did leave the home, just under half of children (46%) did so for longer than 60 minutes and leaving the home environment for this amount of time was found to be important for children being sufficiently active (i.e. MVPA-60minutes guidelines, OR=7.9). The government guidance during the first and all subsequent lockdowns (November 2020, January - currently 2021) has been to minimise the time spent outside of the home, and there has been a common misconception that exercising away from the home should be for no longer than one hour.^45^ This study illustrates the importance of extending the amount of time away from the home for children to be physically active, and if this can be done safely, should be promoted.

The places children most frequently reported going to were, the streets, and parks, and the most frequent activities reported were walking and riding a bike/scooter. The results showed that children who reported going to the shops had reduced odds of being sufficiently active, therefore illustrating that getting out of the home environment to places which are conducive to being active (e.g. streets, parks, greenspaces) should be encouraged, whilst adhering to current COVID-19 guidelines and taking always necessary precautions (e.g. staying 2m apart). Furthermore, as would be expected, children who reported engaging in more vigorous PA such as riding a bike/scooter and playing sports and games were more likely to be sufficiently active than those who reported just walking; suggesting that campaigns should focus on the promotion of these more vigorous types of activities, but also acknowledge that any PA is worthwhile and should be promoted.

There were large differences in whether children reported leaving the home and for how long between ethnic groups. PH and O children left the home significantly less often than their WB peers and for shorter periods. When frequency of leaving the home was controlled for, ethnic PA differences no longer existed between WB and PH children, therefore highlighting an inequality in a key factor for why children were sufficiently active during COVID-19 lockdown (i.e. more PH children stayed at home than WB, therefore were less active). Because of the importance of leaving the home environment to be sufficiently active during COVID-19 lockdown, it is important for policy, strategy and practice to consider why some children were leaving the home environment and why others were not, particularly between different ethnic group. The current study did not directly ask children to report why they had not left their home so this could not be examined. It may have been that those who did not leave the home environment were living in areas less conducive for PA. The following environmental determinants of children and adolescents PA have previously been identified: walkability, availability/access/proximity to recreational facilities, environment aesthetics, negative street characteristics.^46^ All such determinants were not explored in the current study and should be considered in future research to possibly explore the ethnic differences found.

A further influence upon whether children were leaving the home in the current study may have been worries and stress experienced by families during lockdown. Mothers of the children from the sample of this study, who mostly live in areas of high deprivation reported numerous difficulties during the spring 2020 lockdown with many insecurities (financial, employment, housing, clinical symptoms of anxiety and depression) and high levels of anxiety about becoming ill or dying from COVID-19.^47^ Because COVID-19 has disproportionately impacted ethnic minority groups such as Pakistani, South Asian, and Black ethnicities more than White British, with greater ill health and death reported.^47^ Anxiety and fear of ill health and death could be greater within PH and O groups leading to them not wanting to leave the home environment. Negative mainstream media reporting on ethnic minorities violating lockdown protocols and government guidelines, and fear of getting labelled when outside home could have been another reason for ethnic minority children not leaving home during lockdown. More research using qualitative and anthropological methodologies are required to begin to understand this complex phenomenon, because there is a risk of the exacerbation of PA inequalities between ethnic groups, which were well established pre-COVID-19.^4, 16, 23, 48^

The guidance to stay at home during lockdown periods and anxieties surrounding leaving the home, and the association between leaving the house and physical activity, has created a demand for home-based PA interventions for children, with numerous of options being made available.^49, 50^ Previous research on the determinants of childrens home PA are unclear with inconsistent findings.^51^ These programmes which have been developed rapidly may not be evidence based or grounded in behaviour change theory. Moreover, there is a dearth of literature regarding the feasibility, acceptability, efficacy and effectiveness of such home-based PA programmes/interventions.^52^ Home-based PA will likely remain in demand for the foreseeable future as part of the gradual reopening of society, and the changing of the home environment from one promoting mainly sedentary time activities to more physically active activities^51^ is a topic of priority to further understand how best is it for children to be active within their home environments.

A concern of the COVID-19 lockdown(s) has been a possible increase of sedentary (particularly screen behaviours) and disturbances of sleep.^18, 53–55^ Findings from this study showed that the majority of children reported meeting sleep guidelines (9-11 hours a day), engaged in more than one hour a day of TV viewing, playing video games on a console, and doing school work; but also only a small proportion of children (10.1%) reported in meeting ST-guidelines recommendations. Data from Canadian young people also found similar low levels of ST compliance during the first COVID-19 lockdown (11.3%).^19^ But the current study has most likely underestimated the amount of ST, due to the use of a screen for school work (school work item queried any school work, whether using a screen or not) not being factored in the estimate. The high non-compliance of ST recommendations is higher compared to a UK sample of young people pre-COVID (23.1%)^56^, which is unsurprising for children restricted to the home environment for much of their time. Of all of the sedentary ST behaviours, playing a video game on a console for ≥3 hours decreased the odds (OR=0.43-0.52) of children being sufficiently active. This suggests that alongside promotion of leaving the home to support PA during and following the pandemic, reducing the use of sedentary ST, in particular video game usage also needs addressing by public health campaigns. The ethnic and sex differences of sleep and sedentary behaviours (video games, mobile usage, school work) found in this study should be further explored with different outcomes such as educational, emotional and mental health which all have been associated previously with sleep and sedentary behaviours.^57^

The limitations of this study include use of two different child self-reported PA questionnaires (for pre and during COVID) with one questionnaire (amended-YAP) not being formally validated. The BiB study had previously used the validated PAQ-C questionnaire due to availability of children’s questionnaires for previous cohort data collection^25^, before the YAP had been published. In the current study, the PAQ-C was decided by authors not to be suitable for use during lockdown with the majority of children not attending school and being restricted to their homes. A further limitation is that causality of the variables associated with PA-guidelines cannot be implied, and neither can the direction of association, this is due to the cross-sectional nature of data presented. However, the circumstances of COVID-19 and the ability to rapidly survey and receive data from 979 children and continue to follow and collect further data in the future, is a strength of this city-wide cohort study. The ongoing study is providing insights into the lives of children’s and families during an ongoing pandemic and provide scientific insight for policy makers to make evidence informed decisions and guidance.^26, 58^

## Conclusion

The findings of this study are important for practitioners, policy and decision makers to consider in order to begin to understand the impact and consequences that the drastic but required COVID-19 measures (i.e. lockdown) has had upon children’s PA which is a key and vital behaviour for health and development. Key associations have been identified between self-reported PA and the frequency and length of time children went outside of the home. COVID-19 guidelines should factor that many children will not be sufficiently active just in the home environment. Leaving the home for physical activity/exercise for a minimum of 60 minutes, and preferably longer each day safely (staying in household bubbles, social distancing, wearing face coverings where necessary) should be actively prioritised and promoted through campaigns and initiatives. Findings should be considered now during the ongoing COVID-19 crisis to support children’s PA and short-term health and wellbeing; and, once COVID-19 is under control. Policies and interventions to facilitate ‘recovery’ after COVID-19 will be required to prevent potential long-term health problems associated with low levels of PA during the pandemic.

## Data Availability

The datasets used and/or analysed during the specific current study are available from the corresponding author on reasonable request.
Scientists are encouraged and able to use BiB data, which are available through a system of managed open access. The steps below describe how to apply for access to BiB data.
Before you contact BiB, please make sure you have read our Guidance for Collaborators (https://borninbradford.nhs.uk/research/guidance-for-collaborators) Our BiB executive review proposals on a monthly basis and we will endeavor to respond to your request as soon as possible. You can find out about the different datasets which are available (https://borninbradford.nhs.uk/research/documents-data/). If you are unsure if we have the data that you need please contact a member of the BiB team.
Once you have formulated your request please complete the Expression of Interest form available (https://borninbradford.nhs.uk/wp-content/uploads/Expression-of-interest-proforma-v2_DM-12.01.18.doc) and send to
If your request is approved we will ask you to sign a collaboration agreement https://borninbradford.nhs.uk/wp-content/uploads/BiB_CollaborationAgreement-12.01.18.docx and if your request involves biological samples we will ask you to complete a material transfer agreement https://borninbradford.nhs.uk/wp-content/uploads/BiB_CollaborationAgreement-12.01.18.docx.

## List of abbreviations

COVID-19/ COVID: Coronavirus disease 2019
SARS-CoV-2: Severe acute respiratory syndrome coronavirus 2
BiB: Born in Bradford
IMD: Index of multiple deprivation
PA: Physical activity
MVPA: moderate-to-vigorous physical activity
OR: Odds Ratio
ST: Screen time
WB: White British
PH: Pakistani heritage
O: Other
YAP: Youth Activity Profile
PAQ-C: Physical Activity Question – Children

## Declarations

### Ethics approval and consent to participate

Informed consent was gained from parents/carers of children, and children themselves assented to participate. Protocols for the studies was approved by the Health Research Authority and Bradford/Leeds research ethics committee (reference: 16/YH/0320).

### Consent for publication

Not applicable

### Availability of data and materials

The datasets used and/or analysed during the specific current study are available from the corresponding author on reasonable request.

Scientists are encouraged and able to use BiB data, which are available through a system of managed open access. The steps below describe how to apply for access to BiB data.

- Before you contact BiB, please make sure you have read our Guidance for Collaborators (https://borninbradford.nhs.uk/research/guidance-for-collaborators) Our BiB executive review proposals on a monthly basis and we will endeavor to respond to your request as soon as possible. You can find out about the different datasets which are available (https://borninbradford.nhs.uk/research/documents-data/). If you are unsure if we have the data that you need please contact a member of the BiB team.
- nce you have formulated your request please complete the ‘Expression of Interest’ form available (https://borninbradford.nhs.uk/wp-content/uploads/Expression-of-interest-proforma-v2_DM-12.01.18.doc) and send to
- If your request is approved we will ask you to sign a collaboration agreement https://borninbradford.nhs.uk/wp-content/uploads/BiB_CollaborationAgreement-12.01.18.docx and if your request involves biological samples we will ask you to complete a material transfer agreement https://borninbradford.nhs.uk/wp-content/uploads/BiB_CollaborationAgreement-12.01.18.docx.

### Authors’ contributions

DDB wrote the first draft, conducted the study analysis. DDB, ADS and SEB conceived the study aims. DDB, ADS, JH, AS, SAD, SJF, KAS, KLC, MMW, KP, JW, JD designed the questionnaires and decided on the questions included. JH, AS, SAD, KAS, KLC conducted the data collection of the questionnaire. DDB, BK, MA, BH cleaned and prepared data and informed on the analysis. KP, RM, JD designed the data collection protocols and have oversight of the BiB COVID work. All authors aided in the interpretation of the findings and read and approved the final manuscript.

## Acknowledgements

Born in Bradford is only possible because of the enthusiasm and commitment of the Children and Parents in BiB. We are grateful to all the participants, health professionals, schools and researchers who have made Born in Bradford happen.

Authors would like to acknowledge and thank our research interns Emma Young, Isobel Young and Dionysia Markesini for their help and aiding in cleaning data.

## References

1. United Kingdom Government. National lockdown: Stay at Home-Coronavirus cases are rising rapidly across the country. Find out what you can and cannot do, https://www.gov.uk/guidance/national-lockdown-stay-at-home#sports-and-physical-activity (2021).

2. United Kingdom Government. Press Release - PM announces easing of lockdown restrictions: 23 June 2020, https://www.gov.uk/government/news/pm-announces-easing-of-lockdown-restrictions-23-june-2020 (2020).

3. United Kingdom Government. Prime Minister announces new national restrictions-From Thursday, everyone must stay at home, with a limited set of exemptions., https://www.gov.uk/government/news/prime-minister-announces-new-national-restrictions (2020).

4. Sallis JF, Adlakha D, Oyeyemi A, et al. An international physical activity and public health research agenda to inform coronavirus disease-19 policies and practices. J Sport Health Sci 2020: S2095-2546(2020)30064-30068. DOI: 10.1016/j.jshs.2020.05.005.

5. Steene-Johannessen J, Hansen BH, Dalene KE, et al. Variations in accelerometry measured physical activity and sedentary time across Europe – harmonized analyses of 47,497 children and adolescents. International Journal of Behavioral Nutrition and Physical Activity 2020; 17: 38. DOI: 10.1186/s12966-020-00930-x.

6. Guthold R, Stevens GA, Riley LM, et al. Global trends in insufficient physical activity among adolescents: a pooled analysis of 298 population-based surveys with 1&#xb7;6 million participants. The Lancet Child & Adolescent Health 2020; 4: 23–35. DOI: 10.1016/S2352-4642(19)30323-2.

7. Sport England. Active Lives Children and Young People Survey: Academic year 2018/19. 2019.

8. Love R, Adams J, Atkin A, et al. Socioeconomic and ethnic differences in children’s vigorous intensity physical activity: a cross-sectional analysis of the UK Millennium Cohort Study. BMJ Open 2019; 9: e027627. DOI: 10.1136/bmjopen-2018-027627.

9. Eyre ELJ and Duncan MJ. The impact of ethnicity on objectively measured physical activity in children. ISRN Obes 2013; 2013: 757431–757431. DOI: 10.1155/2013/757431.

10. Bailey R, Hillman C, Arent S, et al. Physical activity: an underestimated investment in human capital? J Phys Act Health 2013; 10: 289–308. 2013/04/27. DOI: 10.1123/jpah.10.3.289.

11. Janssen I and Leblanc AG. Systematic review of the health benefits of physical activity and fitness in school-aged children and youth. The international journal of behavioral nutrition and physical activity 2010; 7: 40. 2010/05/13. DOI: 10.1186/1479-5868-7-40.

12. Ekelund U, Luan Ja, Sherar LB, et al. Moderate to Vigorous Physical Activity and Sedentary Time and Cardiometabolic Risk Factors in Children and Adolescents. JAMA 2012; 307: 704–712. DOI: 10.1001/jama.2012.156.

13. Wu XY, Han LH, Zhang JH, et al. The influence of physical activity, sedentary behavior on health-related quality of life among the general population of children and adolescents: A systematic review. PloS one 2017; 12: e0187668–e0187668. DOI: 10.1371/journal.pone.0187668.

14. World Health Organisation (WHO). Global action plan on physical activity 2018-2030: more active people for a healthier world. 2018. Geneva: World Health Organization.

15. Nightingale CM, Rudnicka AR, Owen CG, et al. Patterns of adiposity and obesity among South Asian and white European children: Child Heart and Health Study in England. Journal of Epidemiology and Community Health 2009; 63: 60. DOI: 10.1136/jech.2009.096727h.

16. Van Lancker W and Parolin Z. COVID-19, school closures, and child poverty: a social crisis in the making. The Lancet Public Health 2020; 5: e243–e244. DOI: 10.1016/S2468-2667(20)30084-0.

17. Douglas M, Katikireddi SV, Taulbut M, et al. Mitigating the wider health effects of covid-19 pandemic response. BMJ 2020; 369: m1557. DOI: 10.1136/bmj.m1557.

18. Becker SP and Gregory AM. Editorial Perspective: Perils and promise for child and adolescent sleep and associated psychopathology during the COVID-19 pandemic. Journal of Child Psychology and Psychiatry 2020; 61: 757–759. DOI: https://doi.org/10.1111/jcpp.13278.

19. Moore SA, Faulkner G, Rhodes RE, et al. Impact of the COVID-19 virus outbreak on movement and play behaviours of Canadian children and youth: a national survey. International Journal of Behavioral Nutrition and Physical Activity 2020; 17: 85. DOI: 10.1186/s12966-020-00987-8.

20. Guerrero MD, Vanderloo LM, Rhodes RE, et al. Canadian children’s and youth’s adherence to the 24-h movement guidelines during the COVID-19 pandemic: A decision tree analysis. J Sport Health Sci 2020; 9: 313–321. DOI: https://doi.org/10.1016/j.jshs.2020.06.005.

21. Sport England. Active Lives Children and Young People Survey Coronavirus (Covid19) Report 2021.

22. Stockwell S, Trott M, Tully M, et al. Changes in physical activity and sedentary behaviours from before to during the COVID-19 pandemic lockdown: a systematic review. BMJ Open Sport &amp;amp; Exercise Medicine 2021; 7: e000960. DOI: 10.1136/bmjsem-2020-000960.

23. Berkhout E, Galasso N, Lawson M, et al. The Inequality Virus: Bringing together a world torn apart by coronavirus through a fair, just and sustainable economy. 2021.

24. Wright J, Small N, Raynor P, et al. Cohort Profile: The Born in Bradford multi-ethnic family cohort study. International Journal of Epidemiology 2012; 42: 978–991. DOI: 10.1093/ije/dys112.

25. Bird PK, McEachan RRC, Mon-Williams M, et al. Growing up in Bradford: protocol for the age 7–11 follow up of the Born in Bradford birth cohort. BMC public health 2019; 19: 939. DOI: 10.1186/s12889-019-7222-2.

26. McEachan R, Dickerson J, Bridges S, et al. The Born in Bradford COVID-19 Research Study: Protocol for an adaptive mixed methods research study to gather actionable intelligence on the impact of COVID-19 on health inequalities amongst families living in Bradford [version 1; peer review: 2 approved]. Wellcome Open Research 2020; 5. DOI: 10.12688/wellcomeopenres.16129.1.

27. City of Bradford Metropolitan District Council. About Bradford - Population 2020.

28. Office for National Statistics. 2011 Census: Population and Household Estimates for England and Wales, (2016, March 2011).

29. Dickerson J, Bird PK, McEachan RRC, et al. Born in Bradford’s Better Start: an experimental birth cohort study to evaluate the impact of early life interventions. BMC public health 2016; 16: 711. DOI: 10.1186/s12889-016-3318-0.

30. Ministry of Housing CaLG. English indices of deprivation 2019, (2019, 20/10/2020).

31. Fairclough SJ, Christian DL, Saint-Maurice PF, et al. Calibration and Validation of the Youth Activity Profile as a Physical Activity and Sedentary Behaviour Surveillance Tool for English Youth. Int J Environ Res Public Health 2019; 16 2019/10/05. DOI: 10.3390/ijerph16193711.

32. Saint-Maurice PF and Welk GJ. Validity and Calibration of the Youth Activity Profile. PLOS ONE 2015; 10: e0143949. DOI: 10.1371/journal.pone.0143949.

33. Crocker PR, Bailey DA, Faulkner RA, et al. Measuring general levels of physical activity: preliminary evidence for the Physical Activity Questionnaire for Older Children. Medicine and science in sports and exercise 1997; 29: 1344–1349. 1997/11/05. DOI: 10.1097/00005768-199710000-00011.

34. Tremblay MS, Carson V, Chaput JP, et al. Canadian 24-Hour Movement Guidelines for Children and Youth: An Integration of Physical Activity, Sedentary Behaviour, and Sleep. Appl Physiol Nutr Metab 2016; 41: S311–327. 2016/06/17. DOI: 10.1139/apnm-2016-0151.

35. US Department of Health and Human Services Office of Disease Prevention and Health Promotion. Healthy people 2020. 2011.

36. Moore JB, Hanes JC, Jr., Barbeau P, et al. Validation of the Physical Activity Questionnaire for Older Children in children of different races. Pediatr Exerc Sci 2007; 19: 6–19. 2007/06/08. DOI: 10.1123/pes.19.1.6.

37. Voss C, Dean PH, Gardner RF, et al. Validity and reliability of the Physical Activity Questionnaire for Children (PAQ-C) and Adolescents (PAQ-A) in individuals with congenital heart disease. PLoS One 2017; 12: e0175806. 2017/04/27. DOI: 10.1371/journal.pone.0175806.

38. Dunton GF, Do B and Wang SD. Early effects of the COVID-19 pandemic on physical activity and sedentary behavior in children living in the U.S. BMC public health 2020; 20: 1351. DOI: 10.1186/s12889-020-09429-3.

39. United Kingdom Government. Local COVID alert levels: what you need to know, https://www.gov.uk/guidance/local-covid-alert-levels-what-you-need-to-know (2020).

40. Telama R. Tracking of physical activity from childhood to adulthood: a review. Obes Facts 2009; 2: 187–195. 2010/01/08. DOI: 10.1159/000222244.

41. Biddle SJH, Pearson N, Ross GM, et al. Tracking of sedentary behaviours of young people: A systematic review. Preventive Medicine 2010; 51: 345–351. DOI: https://doi.org/10.1016/j.ypmed.2010.07.018.

42. Hallal PC, Wells JCK, Reichert FF, et al. Early determinants of physical activity in adolescence: prospective birth cohort study. BMJ 2006; 332: 1002–1007. DOI: 10.1136/bmj.38776.434560.7C.

43. Ferreira I, van der Horst K, Wendel-Vos W, et al. Environmental correlates of physical activity in youth - a review and update. Obes Rev 2007; 8: 129–154. 2007/02/16. DOI: 10.1111/j.1467-789X.2006.00264.x.

44. Cleland V, Timperio A, Salmon J, et al. Predictors of time spent outdoors among children: 5-year longitudinal findings. Journal of Epidemiology and Community Health 2010; 64: 400–406. DOI: 10.1136/jech.2009.087460.

45. Bell A. COVID-19: HISTORY SHOWS PUBLIC TREAT ADVICE AS LAW FOR THE COMMON GOOD, (2020).

46. Carlin A. A life course examination of the physical environmental determinants of physical activity behaviour: a “determinants of diet and physical activity” (DEDIPAC) umbrella systematic literature review. PLoS One 2017; 12. DOI: 10.1371/journal.pone.0182083.

47. Dickerson J, Kelly B, Lockyer B, et al. Experiences of lockdown during the Covid-19 pandemic: descriptive findings from a survey of families in the Born in Bradford study [version 1; peer review: awaiting peer review]. Wellcome Open Research 2020; 5. DOI: 10.12688/wellcomeopenres.16317.1.

48. Razai MS, Kankam HKN, Majeed A, et al. Mitigating ethnic disparities in covid-19 and beyond. BMJ 2021; 372: m4921. DOI: 10.1136/bmj.m4921.

49. Criddle C. Coronavirus creates boom in digital fitness, https://www.bbc.co.uk/news/technology-55318822.

50. Skjong I. Kids Exercise Videos to Help Keep Your Family Moving (and Sane), https://www.nytimes.com/wirecutter/blog/best-kids-exercise-videos/ (2020).

51. Maitland C, Stratton G, Foster S, et al. A place for play? The influence of the home physical environment on children’s physical activity and sedentary behaviour. International Journal of Behavioral Nutrition and Physical Activity 2013; 10: 99. DOI: 10.1186/1479-5868-10-99.

52. Messing S, Rütten A, Abu-Omar K, et al. How Can Physical Activity Be Promoted Among Children and Adolescents? A Systematic Review of Reviews Across Settings. Frontiers in Public Health 2019; 7. Review. DOI: 10.3389/fpubh.2019.00055.

53. Vanderloo LM, Carsley S, Aglipay M, et al. Applying Harm Reduction Principles to Address Screen Time in Young Children Amidst the COVID-19 Pandemic. Journal of Developmental & Behavioral Pediatrics 2020; 41.

54. Margaritis I, Houdart S, El Ouadrhiri Y, et al. How to deal with COVID-19 epidemic-related lockdown physical inactivity and sedentary increase in youth? Adaptation of Anses’ benchmarks. Arch Public Health 2020; 78: 52. 2020/06/10. DOI: 10.1186/s13690-020-00432-z.

55. Lee J. Mental health effects of school closures during COVID-19. The Lancet Child & Adolescent Health 2020; 4: 421. DOI: 10.1016/S2352-4642(20)30109-7.

56. Pearson N, Sherar LB and Hamer M. Prevalence and Correlates of Meeting Sleep, Screen-Time, and Physical Activity Guidelines Among Adolescents in the United Kingdom. JAMA Pediatrics 2019; 173: 993–994. DOI: 10.1001/jamapediatrics.2019.2822.

57. Guerrero MD, Barnes JD, Chaput J-P, et al. Screen time and problem behaviors in children: exploring the mediating role of sleep duration. International Journal of Behavioral Nutrition and Physical Activity 2019; 16: 105. DOI: 10.1186/s12966-019-0862-x.

58. Bradford Institite for Health Research (BIHR). COVID-19 Scientific Advisory Group (C-SAG), https://www.bradfordresearch.nhs.uk/c-sag/ (2021).

