## Appendix 1 supplementary material for "Covid-19 lockdown: Ethnic differences in children’s self-reported physical activity and the importance of leaving the home environment. A longitudinal and cross-sectional study from the Born in Bradford birth cohort study" (from another version of this preprint: version 1, 2021-03-01)

| **Appendix 1:** Full model of multivariable logistic regression analysis of factors (demographic, self-reported sleep and sedentary behaviours behavioural, school attendance, frequency and duration of leaving the home environment) with children's self-reported seven day recall of being sufficiently physically active (60 minutes < usually a day) during COVID-19 UK restrictions (April-June 2020). | | | | | | | | | | | | | | | | | | | | | | | | | |
| --- | --- | --- | --- | --- | --- | --- | --- | --- | --- | --- | --- | --- | --- | --- | --- | --- | --- | --- | --- | --- | --- | --- | --- | --- | --- |
|  | **Model 1 (n=946)** | | | | | | | **Model 2 (n=875)** | | | | | **Model 3 (n=868)** | | | | | | **Model 4 (n=602)** | | | | | | |
|  | **z** | **SE** | **OR** | **(95% CI)** | | ***p*** | | **z** | **SE** | **OR** | **95% CI** | ***p*** | **z** | **SE** | **OR** | **95% CI** | ***p*** | | **z** | **SE** | | **OR** | **95% CI** | | ***p*** |
| **Age** (years) | -2.90 | 0.05 | 0.82 | (0.72-0.94) | | **0.00** | | -2.38 | 0.06 | 0.83 | (0.72-0.97) | **0.02** | -2.35 | 0.06 | 0.83 | (0.12-0.97) | 0.02 | | -2.08 | 0.07 | | 0.81 | (0.67-0.97) | | **0.02** |
| **Gender**- Male (Reference) | Ref | | | | |  | | Ref | | | |  | Ref | | | |  | | Ref | | | | | |  |
| Female | -1.44 | 0.12 | 0.81 | | 0.60-1.08 | 0.15 | | -2.72 | 0.11 | 0.63 | (0.45-0.88) | **0.01** | -2.84 | 0.11 | 0.61 | (0.44-0.86) | **0.01** | | -1.23 | | 0.16 | 0.76 | | (0.49-1.12) | 0.16 |
| **Ethnicity -** White British (Reference) | Ref | | | | |  | | Ref | | | |  | Ref | | | |  | | Ref | | | | | |  |
| Pakistani Heritage | -2.38 | 0.12 | 0.64 | | (0.44-0.92) | **0.02** | | -2.20 | 0.13 | 0.62 | (0.41-0.95) | **0.03** | -1.46 | 0.16 | 0.72 | (0.47-1.12) | 0.14 | | -0.66 | | 0.25 | 0.82 | | (0.54-1.59) | 0.79 |
| Other ethnicities | -2.39 | 0.13 | 0.57 | | (0.35-0.90) | **0.02** | | -2.66 | 0.13 | 0.50 | (0.30-0.83) | **0.01** | -2.37 | 0.14 | 0.53 | (0.31-0.90) | **0.02** | | -2.15 | | 0.16 | 0.48 | | (0.31-1.10 | 0.10 |
| **Index of Multiple Deprivation**  Most Deprived (Reference) | Ref | | | | |  | | Ref | | | |  | Ref | | | |  | | Ref | | | | | |  |
| 2nd most deprived | 1.69 | 0.33 | 1.46 | | (0.94-2.28) | 0.09 | | 1.08 | 0.31 | 1.30 | (0.81-2.08) | 0.28 | 0.82 | 0.30 | 1.22 | (0.76-1.98) | 0.41 | | 0.99 | | 0.43 | 1.41 | | (0.78-2.54) | 0.26 |
| 3rd most deprived | -0.11 | 0.22 | 0.97 | | (0.62-1.52) | 0.91 | | -1.02 | 0.19 | 0.78 | (0.48-1.26) | 0.31 | -0.79 | 0.20 | 0.82 | (0.51-1.33) | 0.43 | | -0.98 | | 0.24 | 0.84 | | (0.45-1.57) | 0.58 |
| 4th < most deprived | 1.38 | 0.27 | 1.33 | | (0.89-2.00) | 0.17 | | 0.53 | 0.25 | 1.13 | (0.72-1.75) | 0.60 | 0.48 | 0.26 | 1.12 | (0.71-1.75) | 0.63 | | 0.64 | | 0.35 | 1.40 | | (0.81-2.41) | 0.23 |
| **Attending School** – No (Reference) | Ref | | | | |  | | Ref | | | |  | Ref | | | |  | | Ref | | | | | |  |
| Yes | 1.47 | 0.33 | 1.41 | | (0.89-2.23) | 0.14 | | 1.07 | 0.34 | 1.32 | (0.79-2.18) | 0.29 | 0.56 | 0.31 | 1.16 | (0.69-1.96) | 0.57 | | 0.92 | | 0.42 | 1.33 | | (0.71-2.28) | 0.42 |
| **Meeting Sleep guidelines** Not meeting - less than 9 hours (Reference) |  | | | | | | | Ref | | | |  | Ref | | | |  | | Ref | | | | | |  |
| Yes- meeting guidelines - 9-11 hours |  |  |  |  |  |  |  | 0.85 | 0.44 | 1.32 | (0.69-2.54) | 0.40 | 0.80 | 0.44 | 1.31 | (0.67-2.55) | 0.43 | | 1.07 | | 0.69 | 1.59 | | (0.69-3.55) | 0.28 |
| Sleep more than 11 hours |  |  |  |  |  |  |  | -0.36 | 0.32 | 0.87 | (0.42-1.81) | 0.72 | -0.33 | 0.33 | 0.88 | (0.42-1.13) | 0.74 | | 0.73 | | 0.69 | 1.43 | | (0.51-3.12) | 0.62 |
| **Watching Television**  < 1 hour (Reference) |  |  |  |  |  |  |  | Ref | | | |  | Ref | | | |  | | Ref | | | | | |  |
| 1-3 hours |  |  |  |  |  |  |  | 0.26 | 0.18 | 1.04 | (0.75-1.46) | 0.80 | 0.45 | 0.19 | 1.08 | (0.77-1.52) | 0.65 | | 0.70 | | 0.26 | 1.17 | | (0.82-1.88) | 0.30 |
| 3hr < |  |  |  |  |  |  |  | -1.26 | 0.18 | 0.74 | (0.46-1.19) | 0.21 | -1.47 | 0.17 | 0.69 | (0.43-1.23) | 0.14 | | -1.02 | | 0.22 | 0.74 | | (0.42-1.31) | 0.30 |
| **Video games on a games console** < 1 hour (Reference) |  |  |  |  |  |  |  | Ref | | | |  | Ref | | | |  | | Ref | | | | | |  |
| 1-3 hours |  |  |  |  |  |  |  | -0.49 | 0.17 | 0.91 | (0.63-1.32) | 0.62 | -0.43 | 0.18 | 0.92 | (0.63-1.33) | 0.67 | | -0.26 | | 0.23 | 0.99 | (0.63-1.55) | | 0.95 |
| 3hr < |  |  |  |  |  |  |  | -3.79 | 0.10 | 0.43 | (0.28-0.67) | **0.00** | -3.55 | 0.10 | 0.45 | (0.29-0.70) | **0.00** | | -2.36 | | 0.14 | 0.52 | (0.31-0.89) | | **0.02** |
| **Computers/tablets use for social activity**  < 1 hour (Reference) |  |  |  |  |  |  |  | Ref | | | |  | Ref | | | |  | | Ref | | | | | |  |
| 1-3 hours |  |  |  |  |  |  |  | 0.51 | 0.23 | 1.11 | (0.74-1.68) | 0.61 | 0.36 | 0.23 | 1.08 | (0.71-1.63) | 0.72 | | 1.15 | | 0.35 | 1.34 | (0.88-2.33) | | 0.15 |
| 3hr < |  |  |  |  |  |  |  | -0.74 | 0.24 | 0.43 | (0.44-1.45) | 0.46 | -0.47 | 0.27 | 0.86 | (0.47-0.71) | 0.64 | | 1.19 | | 0.71 | 1.67 | (0.61-3.00) | | 0.46 |
| **Mobile phone use**  < 1 hour (Reference) |  |  |  |  |  |  |  | Ref | | | |  | Ref | | | |  | | Ref | | | | | |  |
| 1-3 hours |  |  |  |  |  |  |  | 0.13 | 0.25 | 1.03 | (0.64-1.66) | 0.90 | 0.42 | 0.27 | 1.11 | (0.69-1.79) | 0.68 | | 0.15 | | 0.32 | 1.04 | (0.56-1.75) | | 0.97 |
| 3hr < |  |  |  |  |  |  |  | -0.97 | 0.23 | 0.73 | (0.39-1.37) | 0.33 | -0.65 | 0.26 | 0.81 | (0.43-1.52) | 0.51 | | -0.75 | | 0.31 | 0.73 | (0.27-1.36) | | 0.23 |
| **School Work**  < 1 hour (Reference) |  |  |  |  |  |  |  | Ref | | | |  | Ref | | | |  | | Ref | | | | | |  |
| 1-3 hours |  |  |  |  |  |  |  | -0.58 | 0.18 | 0.89 | (0.60-1.32) | 0.56 | -0.58 | 0.18 | 0.89 | (0.59-1.32) | 0.56 | | -0.64 | | 0.22 | 0.85 | (0.56-1.34) | | 0.44 |
| 3hr < |  |  |  |  |  |  |  | 0.89 | 0.25 | 1.20 | (0.80-1.80) | 0.37 | 0.86 | 0.25 | 1.20 | (0.79-1.81)) | 0.39 | | -0.11 | | 0.27 | 1.03 | (0.60-1.63) | | 0.97 |
| **Frequency of leaving home** Stayed at Home (Reference) |  |  |  |  |  |  |  |  | | | | | Ref | | | |  | | Ref | | | | | |  |
| Once a day (Reference - Model 3) |  |  |  |  |  |  |  |  |  |  |  |  | 2.16 | 0.33 | 1.57 | (1.04-2.36) | **0.03** | |  |  |  |  |  |  |  |
| More than once a day |  |  |  |  |  |  |  |  |  |  |  |  | 3.97 | 0.69 | 2.73 | (1.66-4.48) | **0.00** | | 0.32 | | 0.26 | 1.08 | (0.73-1.81) | | 0.54 |
| **Duration of time away from home**  < 30 minutes (Reference) |  |  |  |  |  |  |  |  |  |  |  |  |  | | | | | | Ref | | | | | |  |
| 31-60 minutes |  |  |  |  |  |  |  |  |  |  |  |  |  |  |  |  |  |  | 1.99 | | 0.88 | 2.21 | (1.27-5.77) | | **0.01** |
| 60 minutes < |  |  |  |  |  |  |  |  |  |  |  |  |  |  |  |  |  |  | 4.95 | | 3.3 | 7.9 | (4.45-21.53) | | **0.00** |
| **Where did children usually go when leaving the home?**  Street (reference) |  | | | | | | |  | | | | |  | | | | | | Ref | | | | | | |
| Shops |  |  |  |  |  |  |  |  |  |  |  |  |  |  |  |  |  |  | -1.98 | | 0.19 | 0.36 | (0.13-0.98) | | 0.04 |
| Park |  |  |  |  |  |  |  |  |  |  |  |  |  |  |  |  |  |  | 0.36 | | 0.27 | 1.09 | (0.68-1.77) | | 0.72 |
| Greenspace/nature |  |  |  |  |  |  |  |  |  |  |  |  |  |  |  |  |  |  | -0.28 | | 0.33 | 0.97 | (0.49-1.88) | | 0.92 |
| Other neighbourhood areas |  |  |  |  |  |  |  |  |  |  |  |  |  |  |  |  |  |  | -0.10 | | 0.33 | 0.90 | (44.3-1.85) | | 0.78 |
| **Usually do, Walk** - No(reference) |  |  |  |  |  |  |  |  |  |  |  |  |  |  |  |  |  |  | Ref | | | | | | |
| Yes |  |  |  |  |  |  |  |  |  |  |  |  |  |  |  |  |  |  | 0.54 | | 28.9 | 1.15 | (0.70-1.88) | | 0.59 |
| **Usually do, Run/Jog** -  No (reference) |  |  |  |  |  |  |  |  |  |  |  |  |  |  |  |  |  |  | Ref | | | | | | |
| Yes |  |  |  |  |  |  |  |  |  |  |  |  |  |  |  |  |  |  | 3.02 | | 0.53 | 2.13 | (1.30-3.47) | | 0.00 |
| **Usually do, Ride bike/scoot** -  No (reference) |  |  |  |  |  |  |  |  |  |  |  |  |  |  |  |  |  |  | Ref | | | | | | |
| Yes |  |  |  |  |  |  |  |  |  |  |  |  |  |  |  |  |  |  | 1.99 | | 0.32 | 1.52 | (1.01-2.31) | | 0.04 |
| **Usually do, Play, Sports or Games** - No (reference) |  |  |  |  |  |  |  |  |  |  |  |  |  |  |  |  |  |  | Ref | | | | | | |
| Yes |  |  |  |  |  |  |  |  |  |  |  |  |  |  |  |  |  |  | 2.10 | | 0.77 | 2.13 | (1.1-4.31) | | 0.03 |
| **Usually do, Other** - No (reference) |  |  |  |  |  |  |  |  |  |  |  |  |  |  |  |  |  |  | Ref | | | | | | |
| Yes |  |  |  |  |  |  |  |  |  |  |  |  |  |  |  |  |  |  | -0.08 | | 0.51 | 0.96 | (0.34-2.70) | | 0.93 |
| Constant | 1.76 | 2.715117 | 3.67 | | (0.86-15.64) | | 0.078 | 1.68 | 3.88 | 4.39 | (0.78-24.83) | 0.09 | 1.06 | 2.42 | 2.64 | (0.44-15.85) | | 0.29 | -0.32 | | 0.82 | 0.6748692 | (0.06-7.25) | | 0.75 |
| Log liklihood | -539.2033 | | | | | | | -483.34 | | | | | -471.39388 | | | | | | -310.63993 | | | | | | |
| Pseudo r-square | 0.029 | | | | | | | 0.062 | | | | | 0.077 | | | | | | 0.1758 | | | | | | |
| Likelihood-Ratio chi-sqaure (df) | 32.17 (8), p=0.0001 | | | | | | | 63.34 (20), p=.0001 | | | | | 78.85(22), p=.0000 | | | | | | 106.86(32), p=0.000 | | | | | | |
